# The Presence and Nature of AI-Use Disclosure Statements in Medical Education Journals: A bibliometric study

**DOI:** 10.1101/2025.11.11.25340015

**Authors:** M. Ans, L. Maggio, H. Algodi, J. Costello, E. Driessen, K. Oswald, L. Lingard

## Abstract

**Background:** As AI-use becomes more common in research, disclosure policies have emerged to ensure transparency and appropriateness. However, database research in other fields suggests that disclosure may lag behind AI-use. Medical education journal editors report that submitted manuscripts rarely include AI-use disclosures, and they perceive a lack of clarity regarding when and how AI-use should be disclosed. However, we lack objective evidence regarding the incidence and nature of AI-use disclosure in medical education.

**Methods:** Using bibliometric methods, we searched a database of 24 leading medical education journals for articles published between January and July 2025 (n=2,762 articles). Screening with *Covidence* software excluded 716 non-empirical and/or non-English language articles. The remainder (n=2,046) were examined for the presence of AI-use disclosures, which were content-analyzed.

**Results:** 2.5% of empirical articles (n=51) had an AI disclosure statement. *BMC Medical Education* contained the most disclosures (24), followed by *Medical Teacher* (7) and *Journal of Surgical Education* (4). Forty-two articles were authored in non-native English-speaking countries, and 69.4% of all first authors had begun publishing in the past decade. Disclosures averaged 43 words and described use superficially: most commonly “editing” and “translation”. Of 18 named tools, ChatGPT was most common. Most disclosures explicitly attested to author responsibility for AI-produced material. Disclosures usually appeared in acknowledgements; those located in methods lacked responsibility attestation. Negative disclosures attesting that AI was not used were also present.

**Discussion:** AI-use disclosures in medical education journals are rare and appear mostly in work from non-native English-speaking regions of the world. A shared disclosure practice is evident: name the tool and affirm author responsibility, but describe use superficially. This suggests a practice of “safe” disclosure that may be more performative than informative, therefore failing to satisfy the goal of ensuring transparent and ethical AI use in research.

## Background

As AI-use becomes more common in research^1^, scientific communities rely on disclosure to ensure that AI is used ethically and appropriately. Trust in science depends on this^2^, which is why disclosure policies from organizations such as the International Committee of Medical Journal Editors^3^, publishers and journals^4,5^ share an emphasis on transparency and accountability: authors are required to explicitly disclose whether they used AI-assisted technologies in the production of submitted work, and to take responsibility for all AI-produced material^6,7^.

However, emerging research suggests that AI disclosure may be lagging behind AI use. A 2025 Nature survey of 5000 researchers found diverging views of disclosure^8^. For instance, when asked whether they had used AI to write a section of a paper and not disclosed the AI use, 17% of mid-career researchers said ‘Yes’, and 47% said ‘No, but I would be willing to’. Database research also suggests that AI-use outstrips disclosure. For instance, one comparative study of >5 million articles in the Dimensions database reported a 468% increase in clusters of ChatGPT-preferred positive terms (e.g., meticulous, intricate, commendable) in texts published in 2023 after the model became available; only .1% of those papers contained language suggesting disclosure^9^. Similarly, a 2024 analysis of abstracts published in high-impact orthopaedic journals found that of 28 containing AI-generated text, only 1 disclosed AI-use despite journal requirements^10^.

AI-use is a hot topic in contemporary medical education conversations: international conferences are abuzz with keynotes, panels, workshops, and papers related to AI^11^. But while it is increasingly apparent that scholars are using AI ^12^, the nature and pattern of our disclosure practices is less clear. In a recent interview study, medical education journal editors described infrequent experience with AI-use disclosures in submitted manuscripts^13^. Most suspected that medical education researchers might be using AI without disclosing, and worried that this may arise from a lack of clarity regarding when disclosure is necessary and what details are required.

To deepen understanding of AI-use disclosure in medical education, we require objective evidence of researchers’ current disclosure practices. Therefore, this study asks: what is the rate and nature of AI-use disclosure in empirical research papers in medical education journals?

## Methods

We conducted a bibliometric study to identify and characterize AI disclosure statements appearing in medical education research articles. Our sampling frame included all articles published between January 1 to June 30, 2025, in the 24 leading medical education journals listed in the Medical Education Journals List^14^. We excluded non-empirical publications (e.g., commentaries, letters, perspectives) both because we perceived the stakes to be higher for disclosing AI-use in research than in commentaries and we anticipated that the conventional genre of empirical articles would support meaningful comparisons regarding disclosure content and location.

Screening and article selection were managed in *Covidence*, a web-based knowledge synthesis software. MA, HA, JC, and LM independently screened the titles and abstracts of all articles to exclude non-empirical publications. Full-texts of the remaining articles were then examined by MA and HA for AI-use disclosure statements. We classified articles as having an AI statement if they explicitly disclosed the use of an AI tool (e.g., ChatGPT) or described how AI was used in the preparation of the work. Articles that merely focused on, evaluated, or discussed an AI tool, without indicating that AI was used to generate or assist with the manuscript, were not counted as having an AI disclosure statement. For example, an article studying the effectiveness of AI in clinical reasoning, but lacking an explicit disclosure of AI involvement in the authorship process, would not be considered to have a statement. Articles with an AI-use disclosure statement proceeded to data extraction (Figure 1).

**Figure 1.**
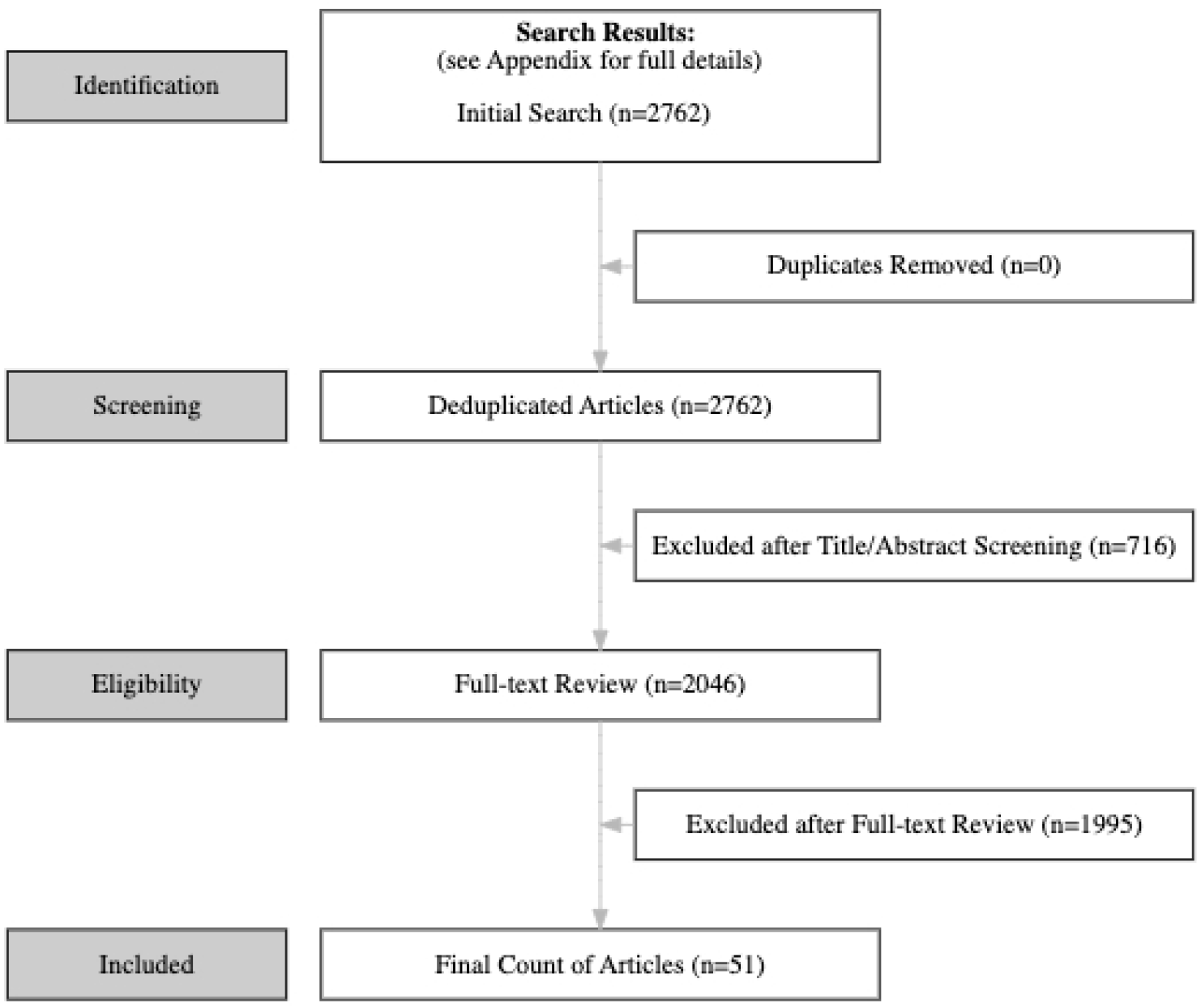
PRISMA Chart

MA and HA each individually extracted data from approximately half of the articles using a structured Google Sheets template. They met regularly to ensure consistent interpretation discussing ambiguous or complex disclosure statements and reaching consensus through discussion. For each article with an AI-use disclosure statement, we extracted:

- Bibliographic variables: DOI, article title, journal name, publication date, journal impact factor, publisher.
- Author-level variables (first author only): name, total publications, total citations, H-index, year of first publication, institutional affiliation, career position, and country of affiliation. These data were collected using Web of Science (WoS). Although we recognize that authors may have publications not indexed in the WoS, we viewed this as a reasonable proxy measure for publishing experience.

For bibliographic and author-level variables, in July 2025, we downloaded the metadata from WoS; if unavailable on WoS, we queried Scopus, except for author career position. Author career position was extracted from the article’s author information or if unavailable, from ResearchGate. Additionally, based on a review of journal and publisher websites, we identified if AI reporting guidance was provided.

Disclosure-specific extractions focused on the name of AI tool(s) used, purpose of AI use, location of the disclosure statement (e.g., methods, acknowledgements, disclosures), verbatim text of the statement, statement word count, and the presence or absence of an attestation of responsibility (e.g., authors remain accountable for the content). We also extracted measures of transparency to situate AI disclosure statements within the broader context of established transparency practices. These included open access status, presence of a funding statement, conflict of interest disclosure, and data availability statement.

For analysis, we conducted descriptive statistics to characterize the bibliographic and author-level variables. To describe the content of AI-use disclosure statements, we conducted a content analysis for AI tool used, the nature of usage, and any reference to responsibility attestation.

## Results

Over the study period, 2,762 articles were published in the medical education journals sampled. Of these, we excluded 716 (25.9%) non-empirical articles. Of the remaining 2,046 articles, 2.5% (n = 51) had an AI disclosure statement.

15,16,17,18,19,20,21,22,23,24,25,26,27,28,29,30,31,32,33,34,35,36,37,38,39,40,41,42,43,44,45,46,47,48,49,50,51,52,53,54,55,56,57,58,59,60,61,62,63,64,65,66

Thirteen of the journals included articles with disclosures. Of these, 12 (92.3%) provided author guidance on disclosure on their website/author instructions, or on their publisher’s website (Table 1). *BMC Medical Education* published nearly half the identified statements (n = 24, 47.1%), followed by *Medical Teacher* (n = 7, 13.7%) and the *Journal of Surgical Education* (n = 4, 7.8%). Nine journals did not include any articles with disclosures. Two journals (*African Journal of Health Professional Education* and *BMJ Stimulation & Technology Enhanced Learning*) did not publish any empirical research during the study period and, therefore, were not examined for AI-use disclosures.

**Table 1.**
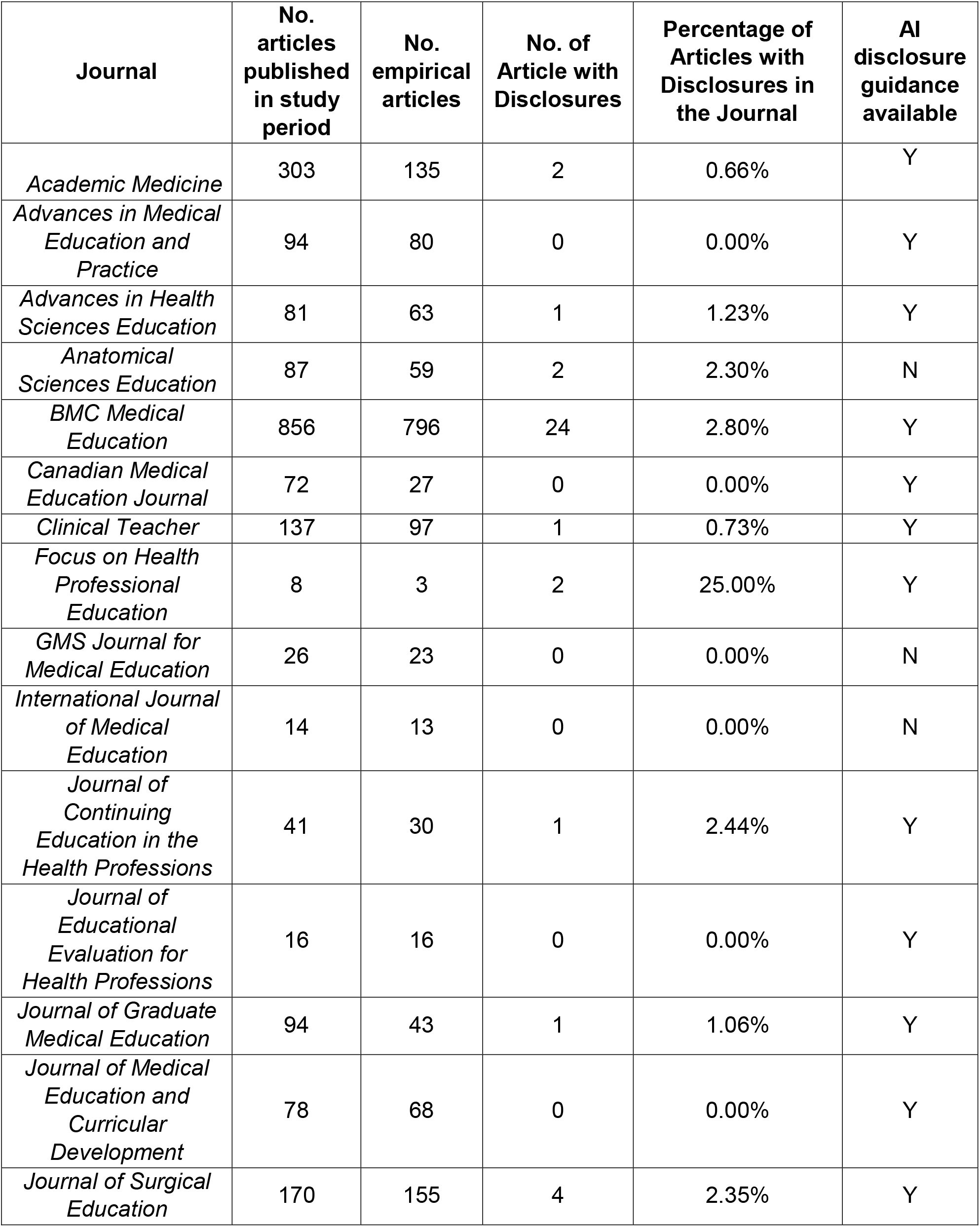
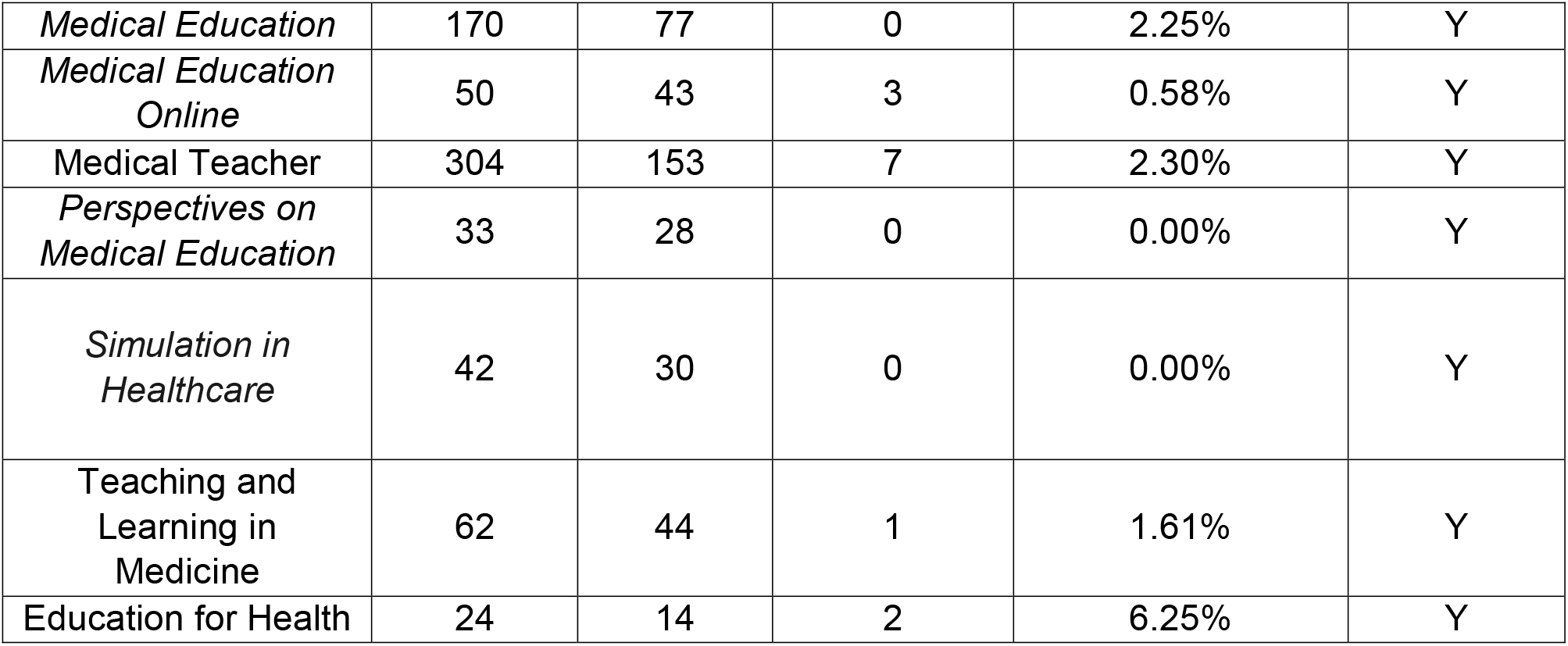
Distribution of AI-Use Disclosures by Journal. *\*\* African Journal of Health Professional Education and BMJ Stimulation & Technology Enhanced Learning* did not have any empirical studies published to analyze.

| **Journal** | **No. articles published in study period** | **No. empirical articles** | **No. of Article with Disclosures** | **Percentage of Articles with Disclosures in the Journal** | **AI disclosure guidance available** |
| --- | --- | --- | --- | --- | --- |
| *Academic Medicine* | 303 | 135 | 2 | 0.66% | Y |
| *Advances in Medical Education and Practice* | 94 | 80 | 0 | 0.00% | Y |
| *Advances in Health Sciences Education* | 81 | 63 | 1 | 1.23% | Y |
| *Anatomical Sciences Education* | 87 | 59 | 2 | 2.30% | N |
| *BMC Medical Education* | 856 | 796 | 24 | 2.80% | Y |
| *Canadian Medical Education Journal* | 72 | 27 | 0 | 0.00% | Y |
| *Clinical Teacher* | 137 | 97 | 1 | 0.73% | Y |
| *Focus on Health Professional Education* | 8 | 3 | 2 | 25.00% | Y |
| *GMS Journal for Medical Education* | 26 | 23 | 0 | 0.00% | N |
| *International Journal of Medical Education* | 14 | 13 | 0 | 0.00% | N |
| *Journal of Continuing Education in the Health Professions* | 41 | 30 | 1 | 2.44% | Y |
| *Journal of Educational Evaluation for Health Professions* | 16 | 16 | 0 | 0.00% | Y |
| *Journal of Graduate Medical Education* | 94 | 43 | 1 | 1.06% | Y |
| *Journal of Medical Education and Curricular Development* | 78 | 68 | 0 | 0.00% | Y |
| *Journal of Surgical Education* | 170 | 155 | 4 | 2.35% | Y |
| *Medical Education* | 170 | 77 | 0 | 2.25% | Y |
| *Medical Education Online* | 50 | 43 | 3 | 0.58% | Y |
| Medical Teacher | 304 | 153 | 7 | 2.30% | Y |
| *Perspectives on Medical Education* | 33 | 28 | 0 | 0.00% | Y |
| *Simulation in Healthcare* | 42 | 30 | 0 | 0.00% | Y |
| Teaching and Learning in Medicine | 62 | 44 | 1 | 1.61% | Y |
| Education for Health | 24 | 14 | 2 | 6.25% | Y |

*Focus on Health Professional Education* had the highest proportion of disclosure statements (25%), although this reflects a small denominator of total articles published, with 2 of 8 containing disclosure statements. Across the other 23 journals, the prevalence of AI-use disclosure statements was low, with no journal exceeding 7% of published articles (Table 1).

### Author Characteristics

Authors were affiliated with institutions across all six World Health Organization (WHO) regions (Table 2) with 24 countries represented (Online Supplemental Appendix A). Fifty unique first authors from 46 institutions included disclosure statements in their articles. Authors affiliated with institutions in the Western-Pacific region accounted for the largest share of disclosures (n = 15, 29.4%), followed by Europe (n = 11, 21.6%) and the Americas (n = 11, 21.6%). Disclosures were also reported by authors from the Mediterranean (n = 9, 17.6%), Southeast Asia (n = 4, 7.8%), and Africa (n = 1, 2.0%). At the country level, USA, Germany, Iran and Australia were most represented.

**Table 2.** Regional Distribution (WHO Regions) of First Authors of Articles with AI-Use Disclosures (n = 51)

| **Region** | **Number of Articles (%)** |
| --- | --- |
| Western Pacific (e.g., Australia, China, South Korea) | 15 (29) |
| Americas (e.g., North, Central, South America) | 11 (22) |
| Europe (e.g., Germany, Netherlands) | 11 (22) |
| Eastern Mediterranean (e.g., Afghanistan, Egypt, Iran) | 9 (18) |
| South-East Asia (e.g., Bhutan, Bangladesh, India) | 4 (8) |
| Africa (e.g., South Africa) | 1 (2) |
| Total | 51 (100) |

On average, first authors had published a median of 11.5 publications (range 1–142, SD=34.7). The median h-index, a citation-based metric, for authors was 4 (range 0-35, SD=7.3) with the median author citations received being 55 (range 0–10,214, SD=2266.2). Thirty-three (66.0%) authors published their first article within the last decade, and for 3 (6.0%) authors, this was their first publication.

Author career positions were variable. Among 51 first authors, there were 50 unique authors with the most common designation being Assistant Professor (n = 9, 17.6%), followed by: other (e.g. clinical roles or administrative positions) (n = 8, 15.7%), trainees or early career researchers (e.g. residents, PhD students, post-doctoral researchers) (n = 7, 13.7%), Associate Professor (n = 6, 11.8%), lecturers/senior lecturers (n = 6, 11.8%),research staff or fellows (n = 6, 11.8%), and Professor (n = 4, 7.8%). Four author career positions were unreported.

### AI tools and Nature of AI-Use

Authors disclosed the use of 18 unique AI tools. ChatGPT was the most disclosed tool (n = 19, 37.3%), followed by Otter.ai (n = 11, 21.6%), and Editage (n = 3, 5.9%) (Table 3). Five articles (9.8%) combined multiple tools, and 4 (7.8%) disclosed AI use without naming a specific tool. Four articles (7.8%) explicitly included negative disclosure statements, stating that no AI tools were used such as: “*Generative AI was not used for any aspect of this study including drafting of the manuscript*.” ^59^ The 4 negative disclosures originated from 2 authors in China, 1 in Hungary, and 1 in Qatar ^46,47,59,62^.

**Table 3.** AI Tools Reported in Disclosure Statements (n = 51) *Some articles reported multiple tools hence the total is greater than 51.

| AI Tool | Brief description of tool | Number of Articles (%)* |
| --- | --- | --- |
| ChatGPT | Large Language Model | 19 (37%) |
| Otter | AI-Powered Transcription Software | 11 (22%) |
| Negative AI Disclosure | N/A | 5 (10%) |
| Unspecified | N/A | 4 (68%) |
| Editage | AI Editing for Researchers | 3 (6%) |
| Claude | AI Assistant | 2 (4%) |
| Curie | Clinical AI Assistant | 2 (4%) |
| ASReview | AI tool for Systematic Reviews | 1 (2%) |
| ClusterBot | AI systematic text categorization | 1 (2%) |
| Consensus.app | AI-powered Search Engine | 1 (2%) |
| Copilot | Generative AI assistant | 2 (2%) |
| DeepL | Comprehensive AI Language Platform | 1 (2%) |
| Grammarly | AI Writing Assistant | 1 (2%) |
| Gemini | Large Language Model | 1 (2%) |
| Hawk | Writing/reading AI Tool | 1 (2%) |
| Open Evidence | Evidence-based clinical decision making AI tool | 1 (2%) |
| POE AI Tools | AI Chatbot Aggregator | 1 (2%) |
| RapidMiner | Data Analytics AI Tool | 1 (2%) |
| Rev.com | Speech to Text Service | 1 (2%) |

We characterized the use of AI into six types, based on the language in the disclosures (Table 4). These categories included: editing, transcription, thematic/data analysis, drafting and article screening. Use of AI for editing purposes was the most reported (n = 28, 54.9%) followed by transcription (n = 12, 23.5%), thematic/data analysis (n = 7, 9.8%), drafting (n = 3, 5.9%), and article screening in knowledge syntheses (n = 2, 3.9%).

**Table 4.** Nature of AI Use in Disclosure Statements (n = 51) ** some articles cited multiple uses of AI hence the total being greater than 49.

| Type of AI Use | Definition | Examples | Number of Articles (%) |
| --- | --- | --- | --- |
| Editing | Polishing | *"Claude v. 3.5 Sonnet has been used for language editing. All content and ideas remain the original work of the authors, with AI assistance to improve linguistic clarity."* | 28 (55) |
| Transcription | Text generation from audio/video | *"We used the Otter.ai transcribing software (Otter.ai Inc, California), with the RA reviewing each transcript for accuracy."* | 12 (24) |
| Analysis | Thematic and Statistical Analysis | *"The authors acknowledge availing of a language quality checker and editing tool that used Curie's AI software freely available from Springer Nature webpage as mentioned in the submission guidelines. The authors also acknowledge the use of Claude 3.5 Sonnet software for thematic analysis of the qualitative data."* | 7 (14) |
| Drafting | Text generation | *"We acknowledge the assistance of the GPT-4 AI language model by OpenAI for support with statistical analysis and drafting of this manuscript"* | 3 (6) |
| Literature Review: Article Screening | Includes article screening as a component of literature review | *"We used an open-source artificial intelligence (AI) tool, ASReview (V.1.4) [22]. ASReview employs a machine learning algorithm that prioritizes articles based on their textual proximity to previously identified relevant articles (by the researchers). The tool consequently reduces the time and effort required during the initial screening phase but does not replace the initial screening of articles by researchers."* | 2 (4) |

AI was most frequently described as being used for language-related functions. Editing was cited in 28 (54.9%) articles, with representative phrasings such as “*we would like to thank Editage for English language editing*” ^40^ or “*the authors used ChatGPT to improve language and readability*.” ^42^ Transcription was described in 12 (23.5%) articles: e.g., “*the interview data was transcribed using Otter*.*ai and then manually checked for accuracy*.” ^63^

Eight unique articles (15.7%) disclosed more substantive AI uses. For example, 4 articles (7.8%) disclosed use of thematic analysis (e.g., one group of “*authors acknowledge the use of Claude 3*.*5 Sonnett for thematic analysis of qualitative data*”).^15^ AI use for data analysis was reported in 4 (7.8%) articles, described in one study as “*ChatGPT was used to assist in identifying descriptive trends and generating narrative summaries*. “^65^ Two articles (3.9%) reported AI for screening purposes, with one article saying, “*We used an open-source artificial intelligence (AI) tool, ASReview (V*.*1*.*4)…ASReview employs a machine learning algorithm that prioritizes articles based on their textual proximity to previously identified relevant articles (by the researchers)*.”^21^

Disclosure statements were typically concise, a single sentence averaging 43 words (Range 11-200, SD 36.1). For instance, one stated that “*DeeplL AI writing assistant was used to enhance this manuscript for methodology, clarity, and style*”^17^, while another reported that “*Chat-GPT 3 was used to improve the clarity of the manuscript and for language editing*”^38^. As these representative examples illustrate, disclosures tended to include general terms such as “enhance” or “improve” without detailing the particular characteristics that were enhanced or how the AI was used to achieve such enhancement. In contrast, only 2 articles (3.9%) had a disclosure word count of over 100 words and offered more precise indications of the nature of AI use. For example, one disclosure stated:

> “*The questionnaire for this study was designed with the assistance of AI tools, demonstrating AI’s potential as a research collaborator. The collected data were initially explored using ChatGPT, an AI-powered language model, to assist in identifying descriptive trends and generating narrative summaries. ChatGPT was not used for statistical calculations or hypothesis testing. Artificial Intelligence tool, specifically OpenAI’s ChatGPT (version GPT-4, accessed via ChatGPT Plus) we used at various stages of this research. The following contributions were made: Questionnaire Development: ChatGPT was used to draft survey items aligned with the research objectives. Prompts included, for example: “Design a student survey to evaluate the use of AI tools in medical education across academic, clinical, and research contexts*.*” Thematic Analysis Assistance: For qualitative responses, ChatGPT helped group answers into initial themes. These were reviewed, corrected, and finalized by the authors to ensure accuracy and context. Narrative Drafting: ChatGPT was used to generate narrative summaries of the findings and to draft portions of the introduction, results, and discussion. Prompts included: “Summarize Likert-scale findings and interpret trends,” and “Rewrite this paragraph in an academic tone*.*” Language Polishing: ChatGPT helped improve the clarity, grammar, and coherence of the manuscript*.*”*^*65*^

Only 29% (n=15) of disclosures included an attestation of responsibility. For example, authors wrote “*After using this tool, the authors reviewed and edited the content as needed and took full responsibility for the content of the publication*.*”*^*25*^

### Location of disclosure statements

Placement of disclosure statements varied. Approximately one-third of the disclosures (n = 19, 37.3%) appeared in the acknowledgements section. Another 15 (29.4%) articles appeared in a dedicated AI-use disclosure section, which consistently appeared at the end of the published manuscript, grouped with other declarations such as conflict of interest or data availability statements. For the remaining statements, 14 (27.5%) articles exclusively reported their disclosure in the methods, 2 (%) in both the methods and a dedicated AI disclosure section, and 1 (%) in the discussion. We noted a relationship between location and content of disclosures: those located in the methods primarily described AI use for transcription, e.g., “*All the audio-recorded interviews were transcribed verbatim into Microsoft Office using Otter*.*ai*.*”*^*39*^

Additionally, attestation statements were more likely to be included in disclosures placed in the acknowledgements or a dedicated AI use disclosure section such as: “*During the preparation of this manuscript, the authors utilized ChatGPT to help refine the language. After using this tool, the authors reviewed and made any necessary adjustments and took full responsibility for the content of the publication*.”^59^ By contrast, these attestation statements were absent when the disclosures appeared in the methods.

### Transparency metrics

We found that 42 articles (82.4%) were published as open-access, 42 articles (82.4%) included a funding statement, 40 articles (78.4%) had a conflict-of-interest statement, and 37 articles (72.5%) had a data availability statement. In most cases, AI-use disclosures appeared directly alongside these established measures of academic transparency (n = 34, 66.7%) at the end of the publication.

## Discussion

Our findings highlight three primary issues for consideration in our field: the low rate of disclosure, the geographical pattern of disclosures concentrated in non-native English-speaking regions, and the practice of superficial, safe disclosures.

Our analysis suggests that AI-use disclosure is rare in medical education: only 2.5% of empirical articles published in 24 medical education journals in the first half of 2025 included a disclosure. This corroborates medical education journal editors’ subjective experience of infrequently seeing disclosures in submitted manuscripts^67^. It also resembles bibliometric results from other research domains: e.g., an analysis of academic radiology articles in 2024 found that 34 of 1998 manuscripts (1.7%) disclosed AI use^68^. Does this low rate signal low AI-use, or a tendency towards nondisclosure of AI-use? Unfortunately, evidence leans towards the latter^69^. For instance, since 2023, analyses by “integrity specialists” have flagged hundreds of published research papers with obvious signs of undisclosed AI use, such as those that contain the phrase “regenerate response”^70^. Given that the field of medical education research is not immune to undisclosed AI-use^71^, we need to explore the underpinning influences.

One influence is the lack of clarity. Not only are journal policies on AI-use and disclosure shifting quickly due to the fast-changing AI landscape, journals’ positions vary. For authors submitting, and resubmitting, their manuscripts, disclosure requirements may appear murky and complicated. Another influence is the divergent attitudes among researchers about AI-use and disclosure: surveys have clearly shown that we don’t agree about what AI should be used for and when those uses require disclosure^8,12^. A third influence is psychological/cultural and institutional barriers that make researchers hesitant to disclose AI-use to improve their writing. They may experience a sense of “demonization” of AI^72^, inducing guilt or shame about using it to support their work^73^ and fear of consequences such as academic stigma and evaluation bias^74^. Finally, the lack of published disclosures may act like a self-fulfilling prophecy: if we suspect others are using AI but we rarely see AI-use disclosures, that can send a tacit message that disclosure is unnecessary.

Our findings also demonstrate a particular geographical pattern to AI-use disclosure in the analyzed articles, with most authored by individuals from non-native English-speaking regions of the world. This likely explains the dominant use of AI tools for language-related tasks, like editing, translating and grammar checking, all of which are valuable to authors writing in English as a non-native language, and resonates with other studies that have reported higher AI-use for writing support by non-native English researchers^75^. Given this geographical pattern of disclosure, medical education needs to be aware of the possibility of stigmatization, given that non-native English academic writing is more likely to raise reviewers’ suspicions about AI-use^76^, and is also known to be prone to misclassification by GPT detectors^77^. The four negative disclosures in our dataset all came from authors in non-native English-speaking regions, which may suggest an attempt to deflect such potential stigmatization. Medical educators need to be aware that, rather than levelling the global playing field in academic publishing, AI-use disclosure might intensify inequity and bias against authors from some parts of the world^78^, further exacerbating the global north dominance in the field’s literature^79,80^.

The disclosures we analyzed were almost uniformly brief, describing general uses such as “editing”, and lacking in detail that would allow a reader to understand precisely how the author engaged with the AI, iteratively refined and structured its work, and verified its outputs. We interpret this as “safe” disclosure practice: authors are mostly including the required elements of tool, task, and attestation, but rarely disclosing substantive, intellectual tasks, or elaborating the ‘how’ of their interaction with the AI. On the one hand, this might be good news: the disclosed uses we found mostly map onto the “acceptable” uses outlined in a recent framework distinguishing ethically acceptable, contingent and suspect uses of AI^81^. On the other hand, though, we worry that our dataset includes few disclosures in the framework’s “contingent” or “suspect” ranges not because researchers are not using AI these ways, but because they perceive themselves to be on shaky ethical ground when they do. While such frameworks can be powerful aids to scholarly conversation about AI use and disclosure, we must take care that they do not unintentionally drive a tacit culture of nondisclosure.

The combination of low disclosure rates and safe disclosure practice produces a “transparency paradox”^82^: mandatory disclosure that does not attend to social complexities leads to both nondisclosed AI-use and disclosure “theatre” in which published disclosures are more performative than informative^83^. One possible solution is for journals to create dropdown menus of possible AI uses and verification strategies, signaling sanctioned uses and expected levels of detail. These menus might draw from emerging disclosure frameworks specifying tiers of usage^84^ or using CREDiT-like author contribution frameworks^85^. Another solution might be to shift away from ‘disclosure’ language altogether and make AI use a routine part of methods reporting. This could help to normalize writing about AI use in manuscripts, thus reducing the negative valence associated with disclosure in the current literature. Given our results, however, we anticipate that authors will need explicit guidance to include responsibility attestations when reporting AI-use within their Methods.

### Limitations

This study has several limitations. First, our analysis was limited to the first 6 months of 2025. Because editors have reported a recent increase in AI-related disclosures^67^, it is possible that disclosure rates were higher during this period, but we lack data to examine changes or trends over time. Moreover, it is possible that the low frequency of disclosures we observed may reflect the field’s publication timelines, given an average lag of approximately 188 days from submission to publication^86^, suggesting that the manuscripts referenced by editors may not yet have been published. Second, we did not compare articles with AI disclosures to those without, which limits our ability to assess differences in overall transparency practices. Finally, because many journals impose word limits, authors may have a limited word count to include and/or elaborate on their AI use, potentially constraining the detail and desire to include a disclosure statement.

## Conclusion

AI-use disclosures in medical education journals are rare and appear mostly in work from non-native English-speaking regions of the world. A shared disclosure practice is evident: name the tool and affirm author responsibility, but describe use superficially. This suggests a practice of safe disclosure that may fail to satisfy the goal of ensuring transparent and ethical AI use in research.

## Data Availability

All data produced in the present work are contained in the manuscript.

## Acknowledgements

This project was funded by the Office of Continuing Professional Development, Schulich School of Medicine & Dentistry, Western University in the form of a 2024 Digital Research & Innovation Grant, as well as by a Summer Research Training Program grant to support the activities of Muhammad Ans.

## Ethics Approval

As a bibliometric study using published documents and no human subjects, this project did not require REB approval.

## References

1. Weixin Liang, Yaohui Zhang, Zhengxuan Wu, et al. 2024. Mapping the Increasing Use of LLMs in Scientific Papers. 2404.01268

2. Wolfgang Blau et al., “Protecting Scientific Integrity in an Age of Generative AI,” Proceedings of the National Academy of Sciences 121, no. 22 (May 28, 2024): 1–2, 10.1073/pnas.2407886121.

3. Recommendations for the Conduct, Reporting, Editing, and Publication of Scholarly Work in Medical Journals. Updated January 2024. https://www.icmje.org/news-and-editorials/updated_recommendations_jan2024.html

4. Eva KW. Medical Education and artificial intelligence: Responsible and effective practice requires human oversight. Med Educ. 2024 Nov;58(11):1260–1261. doi: 10.1111/medu.15495.

5. Flanagin A, Pirracchio R, Khera R, Berkwits M, Hswen Y, Bibbins-Domingo K. Reporting Use of AI in Research and Scholarly Publication—JAMA Network Guidance. JAMA. 2024;331(13):1096–1098. doi:10.1001/jama.2024.3471

6. Ganjavi C, Eppler M B, Pekcan A, Biedermann B, Abreu A, Collins G S et al. Publishers’ and journals’ instructions to authors on use of generative artificial intelligence in academic and scientific publishing: bibliometric analysis BMJ 2024; 384 :e077192 doi:10.1136/bmj-2023-077192

7. Perkins M, Roe J. Academic publisher guidelines on AI usage: A ChatGPT supported thematic analysis. F1000Res. 2024 Jan 16;12:1398.

8. Kwon, Diane. Is it OK for AI to write science papers? Nature survey shows researchers are split. Nature May 2025

9. Gray, Andrew. “ChatGPT” contamination”: estimating the prevalence of LLMs in the scholarly literature.” arXiv preprint 2403.16887 (2024).

10. Pesante, Benjamin & Mauffrey, Cyril & Parry, Joshua. (2024). Rise of the Machines: The Prevalence and Disclosure of Artificial Intelligence-Generated Text in High-Impact Orthopaedic Journals. The Journal of the American Academy of Orthopaedic Surgeons. 32. 10.5435/JAAOS-D-24-00318.

11. The International Association for Health Professions Education, 2025 Conference Program. https://amee.org/amee-2025-archive/amee-2025-programme-2/overview/

12. Kwon, D. Science sleuths flag hundreds of papers that use AI without disclosing it. Nature 641, 290–291 (2025). doi: 10.1038/d41586-025-01180-2

13. L Lingard, E Driessen, K Oswald. (2025). The blurred threshold of AI-use disclosure: International journal editors’ expectations of sufficiency and necessity. doi: 10.1101/2025.07.17.25331725

14. Maggio LA, Ninkov A, Frank JR, Costello JA, Artino AR Jr. Delineating the field of medical education: Bibliometric research approach(es). Med Educ. 2022; 56(4): 387–394. doi:10.1111/medu.14677

15. Atreya A, Gnawali L, Menezes RG, Nepal S. Enhancing medical education in Nepal through problem-based learning (Pbl) and collaborative action research strategies. BMC Med Educ. 2025 May 10;25(1):689.

16. Atreya A, Rajbanshi R, Menezes RG, Acharya A. Evaluation of undergraduate forensic medicine education in Nepal: a critical analysis using Schwab’s five commonplaces and Schubert’s curriculum images. BMC Med Educ. 2025 Jan 29;25(1):147.

17. Baris M, Schaper NV, Weis HS, Fröhlich K, Rustenbach C, Herrmann-Werner A, et al. Surgical simulation in emergency management and communication improves performance, confidence, and patient safety in medical students. Medical Education Online. 2025 Dec 31;30(1):2486976.

18. Bawazeer GA, Alqifari SF, Alkhudair NA, Aldemerdash AF, Aljuffali LA, Korayem GB. The 2024 saudi pharmacotherapy didactic curriculum toolkit: a modified delphi study. BMC Med Educ. 2025 Apr 21;25(1):583.

19. Behmanesh D, Jalilian S, Heydarabadi AB, Ahmadi M, Khajeali N. The impact of hidden curriculum factors on professional adaptability. BMC Med Educ. 2025 Feb 5;25(1):186.

20. Bereuter JP, Geissler ME, Klimova A, Steiner RP, Pfeiffer K, Kolbinger FR, et al. Benchmarking vision capabilities of large language models in surgical examination questions. Journal of Surgical Education. 2025 Apr;82(4):103442.

21. Bock LA, Vaassen S, Van Mook WNKA, Noben CYG. Understanding healthcare efficiency—an AI-supported narrative review of diverse terminologies used. BMC Med Educ. 2025 Mar 20;25(1):408.

22. Bourke J, Caldwell J, Martin R, Grainger R. The need for active allies: A narrative analysis of disabled medical students’ perspectives of their medical school. FoHPE. 2025 Mar 31;26(1):40–59.

23. Cormack CJ, Childs J, Kent F. Sonographer experiences of interprofessional ultrasound education: a qualitative study. J Contin Educ Health Prof [Internet]. 2025 Jan 22 [cited 2025 Nov 5]; Available from: https://journals.lww.com/10.1097/CEH.0000000000000593

24. Darici D, Sieg L, Eismann H, Karsten J. Leader-follower dynamics in medical training: A dual mobile eye-tracking analysis of teacher-student gaze patterns. Medical Teacher. 2025 June 16;1–9.

25. De Almeida Rodrigues DDS, De Almeida MAS, Da Silva Ferreira NC, Alves LA. Anesthesiology competencies in undergraduate medical education: a comparative study of curriculum frameworks in Brazil, Spain, and the United Kingdom. BMC Med Educ. 2025 Apr 11;25(1):516.

26. Derakhshan HB, Shoorei H, Hassanzadeh G, Mansoorian MR, Moshari J, Sajjadi M, et al. Production and educational value of anatomical megamoulages. BMC Med Educ. 2025 Apr 4;25(1):485.

27. Dhillon S, Roque MI, Maylott P, Brooks D, Wojkowski S. Strategies to increase accessibility for students with disabilities in health professional education programs: A scoping review: BEME Review No. 94. Medical Teacher. 2025 July 3;47(7):1062–82.

28. El-Beshbishi SN, Abdel Razek M, El-Marsafawy HM, Hamed O. Use of AI machine learning algorithms to assess medical student engagement and predict performance. EfH. 2025 June 25;38(2):132–44.

29. Elliott AP, Croom T, Watts BV, Horstman MJ, Godwin KM. Curriculum mapping: Visualizing curricular alignment in a competency-based interprofessional fellowship program. Medical Teacher. 2025 Aug 3;47(8):1394–8.

30. Elster MJ, Parsons AS, Collins S, Gusic ME, Hauer KE. ‘We’re like Spider-Man; with great power comes great responsibility’: Coaches’ experiences supporting struggling medical students. Medical Teacher. 2025 Feb;47(2):329–37.

31. Flock C, Boekels R, Herrmann A, Beig I, Lamkemeyer L, Friederich HC, et al. Final year medical students’ expectations for medical education on climate change and planetary health – a qualitative study. Medical Education Online. 2025 Dec 31;30(1):2477670.

32. Gellisch M, Cramer J, Trenkel J, Bäker F, Bablok M, Morosan-Puopolo G, et al. Introducing the contextual digital divide: Insights from microscopic anatomy on usage behavior and effectiveness of digital versus face-to-face learning. Anatomical Sciences Ed. 2025 Apr;18(4):347–64.

33. Ghasemian A, Salehi M, Ghavami V, Yari M, Tabatabaee SS, Moghri J. Exploring dental students’ attitudes and perceptions toward artificial intelligence in dentistry in Iran. BMC Med Educ. 2025 May 19;25(1):725.

34. Gin BC, LaForge K, Burk-Rafel J, Boscardin CK. Macy foundation innovation report part ii: from hype to reality: innovators’ visions for navigating ai integration challenges in medical education. Acad Med. 2025 Sept;100(9S):S22–9.

35. Godlimpi L, Nomatshila SC, Nanjoh MK, Mnyaka OR, Chitha WW, Mabunda SA, et al. Health professionals’ perceptions of the Walter Sisulu University’s integrated longitudinal clinical clerkship on service delivery in rural district hospitals in Eastern Cape Province, South Africa. BMC Med Educ. 2025 Mar 11;25(1):365.

36. Grille S, Boada M, Guillermo C. Advancing hematological cytology education through virtual training. EfH. 2025 June 25;38(2):145–53.

37. Gümüş A, Zengi O, Kazezoğlu C, Uçar KT, Coşkun C, Tek S, et al. Assessment of centrifugation knowledge among medical laboratory personnel: a survey-based study. BMC Med Educ. 2025 Mar 19;25(1):398.

38. Hasan MJ, Islam S, Sujon H, Chowdhury FI, Islam M, Ahmed M, et al. Research involvement among undergraduate medical students in Bangladesh: a multicenter cross-sectional study. BMC Med Educ. 2025 Jan 25;25(1):126.

39. Imwattana K, Ngamskulrungroj P. Comparison of online quizzes and standard summative examination for the evaluation and guidance of students in medical bacteriology and mycology. BMC Med Educ. 2025 Mar 5;25(1):342.

40. Ishido K, Kiriyama K, Poudel S, Hiradate M, Kono Y, Kurashima Y, et al. Differences in preoperative preparation between novice surgeons and experts: a scoping review. Journal of Surgical Education. 2025 July;82(7):103540.

41. Jamieson J, Palermo C, Hay M, Bacon R, Lutze J, Gibson S. An evaluation of programmatic assessment across health professions education using contribution analysis. Adv in Health Sci Educ [Internet]. 2025 June 4 [cited 2025 Nov 5]; Available from: https://link.springer.com/10.1007/s10459-025-10444-5

42. Janatolmakan M, Piri S, Nouri MA, Khatony A. Empowering nursing students: understanding and addressing bullying experiences in clinical training. BMC Med Educ. 2025 Feb 6;25(1):192.

43. Kataoka H, Tokinobu A, Fujii C, Watanabe M, Obika M. Effectiveness of professional-identity-formation and clinical communication-skills programs on medical students’ empathy in the COVID-19 context: comparison between pre-pandemic in-person classes and during-pandemic online classes. BMC Med Educ. 2025 Jan 9;25(1):39.

44. Keshmiri F. The experiences of unprofessionalism among students in dental education: a qualitative study. BMC Med Educ. 2025 Jan 6;25(1):24.

45. Kiger ME, Meyer HS. Ownership of patient care: medical students’ expectations experiences, and evolutions across the core clerkship curriculum. Teaching and Learning in Medicine. 2025 May 27;37(3):287–99.

46. Kiss H, Pikó BF. Risk and protective factors of student burnout among medical students: a multivariate analysis. BMC Med Educ. 2025 Mar 15;25(1):386.

47. Liu S, Zhao Z, Chen X, Chi Y, Yuan S, Cai F, et al. Evaluation of health care providers’ ability to identify patient-ventilator triggering asynchrony in intensive care unit: a translational observational study in China. BMC Med Educ. 2025 Feb 4;25(1):182.

48. Madgwick J, Anderson L, Cornwall J. Supporting minority cultures during initial engagements with body donors in the dissecting room: A pilot study exploring perspectives of Pasifika medical students around culture and cultural safety. Anatomical Sciences Ed. 2025 Feb;18(2):160–71.

49. Matthews K, Barker L, Bourne E, Dixon K, Palermo C. Allied health student placements in residential aged care: attitudes, experiences and impact. The Clinical Teacher. 2025 Apr;22(2):e70041.

50. Meinlschmidt G, Koc S, Boerner E, Tegethoff M, Simacek T, Schirmer L, et al. Enhancing professional communication training in higher education through artificial intelligence(Ai)-integrated exercises: study protocol for a randomised controlled trial. BMC Med Educ. 2025 May 30;25(1):804.

51. Naito T, Hashizumi A, Sakai M, Yamamura E, Iwase M, Yamada K, et al. Sustained effects of bladder point-of-care ultrasound simulation exercise on nursing students: A prospective cohort study. BMC Med Educ. 2025 Jan 25;25(1):127.

52. Ng O, Tay ZH, Chee DZY, Hui Min Lau S, Liu Z, Cleland J, et al. User-centred curriculum mapping: A human-AI hybrid approach. Medical Teacher. 2025 Apr 15;1–3.

53. Quaintance JL, Ngo TL, Wenrich MD, Hatem D, Keeley MG, Lewis JM, et al. Competence, mattering and belonging: An evidence-based and practical approach to understanding and fostering medical student professional identity formation. Medical Teacher. 2025 May 6;1–12.

54. Rahmanti AR, Yang HC, Huang CW, Huang CT, Lazuardi L, Lin CW, et al. Validating nonverbal cues for assessing physician empathy in telemedicine: a Delphi study. Medical Education Online. 2025 Dec 31;30(1):2497328.

55. Rahmanti AR, Yang HC, Huang CW, Huang CT, Lazuardi L, Lin CW, et al. Validating nonverbal cues for assessing physician empathy in telemedicine: a Delphi study. Medical Education Online. 2025 Dec 31;30(1):2497328.

56. Rissmiller B, Thammasitboon S, Benjamin J. Revitalizing scientific poster sessions: a gamification approach at an academic conference. Acad Med. 2025 Oct;100(10):1158–62.

57. Roshal JA, Lund S, L’Huillier JC, Silvestri C, Woodward JM, Gan C, et al. Out of touch: a nationwide mixed-methods e-learning needs assessment of general surgery residents. Journal of Surgical Education. 2025 June;82(6):103514.

58. Saad S, Ali S. Academic resilience in medical students: exploring students’ perception of social support provided by peers and teachers to help at-risk students for the successful academic journey. BMC Med Educ. 2025 Feb 19;25(1):271.

59. Seed Ahmed M, Soltani A, Zahra D, Allouch S, Al Saady RM, Nasr A, et al. Remote online learning reimagined: perceptions and experiences of medical students in a post-pandemic world. BMC Med Educ. 2025 Feb 10;25(1):215.

60. Taoube L, Roberts C, Burgess A, Khanna P, Schneider CR. Navigating the educational landscape in primary care: Medical student interprofessional placements across communities of practice. Medical Teacher. 2025 June 25;1–9.

61. Topf C, Buzo M, Hahlweg P, Scholl I. Medical students’ attitudes toward providing patients with audio recordings of their medical encounters: a cross-sectional online survey. BMC Med Educ. 2025 June 19;25(1):853.

62. Tricard T, Pan J, Song Q, Lu Y, Ye G, Bian Z. Validation of the objective structured assessment of technical skill (Osats) in china. Journal of Surgical Education. 2025 Jan;82(1):103304.

63. Van Heerden C, Hawley M, Jayawardena N, Gray A. Perceptions of feedback up to senior doctors and nurses in a tertiary paediatric hospital: A mixed-methods study. FoHPE. 2025 Mar 31;26(1):20–39.

64. Winkel AF, Burk-Rafel J, Terhune K, Garibaldi BT, DeWaters AL, Co JPT, et al. Large language model– augmented strategic analysis of innovation projects in graduate medical education. Journal of Graduate Medical Education. 2025 May;17(2s):6–9.

65. Yousef M, Deeb S, Alhashlamon K. AI usage among medical students in Palestine: a cross-sectional study and demonstration of AI-assisted research workflows. BMC Med Educ. 2025 May 12;25(1):693.

66. Zhang Z, Liu Y, Sharif-Nia H. Validating the revised plagiarism attitude scale among Malaysian medical sciences students: a psychometric study in multilingual contexts. BMC Med Educ. 2025 Apr 16;25(1):552.

67. L. Lingard, E. Driessen, K. Oswald. The blurred threshold of AI-use disclosure: International journal editors’ expectations of sufficiency and necessity. medRxiv 2025.07.17.25331725; doi: 10.1101/2025.07.17.25331725

68. Jonah Barrett D, Heng R, Perchik JD. Documenting Disclosure: Limited Reporting of Generative AI Usage in Radiology Research Manuscripts. Acad Radiol. 2025 Oct;32(10):5686–5692. doi: 10.1016/j.acra.2025.06.057.

69. Glynn, Alex. “Suspected Undeclared Use of Artificial Intelligence in the Academic Literature: An Analysis of the Academ-AI Dataset.” ArXiv abs/2411.15218 (2024)

70. Kwon D. (2025) Hundreds of papers use AI without Disclosure. Nature 290, Vol 641, 290–291.

71. Masters, Ken. “Medical Teacher’s first ChatGPT’s referencing hallucinations: Lessons for editors, reviewers, and teachers.” Medical Teacher 45.7 (2023): 673–675.

72. Fiorillo L. Confronting the demonization of AI writing: reevaluating its role in upholding scientific integrity. Oral Oncol Rep. 2024;12:100685. doi:10.1016/j.oor.2024.100685

73. Giray L. AI Shaming: The Silent Stigma among Academic Writers and Researchers. Ann Biomed Eng. 2024 Sep;52(9):2319–2324. doi: 10.1007/s10439-024-03582-1. Epub 2024 Jul 8. PMID: 38977530.

74. Ahmed S BaHammam (2025) The Transparency Paradox: Why Researchers Avoid Disclosing AI Assistance in Scientific Writing, Nature and Science of Sleep,, 2569–2574, DOI: 10.2147/NSS.S568375

75. Jonah Barrett D, Heng R, Perchik JD. Documenting Disclosure: Limited Reporting of Generative AI Usage in Radiology Research Manuscripts. Acad Radiol. 2025 Oct;32(10):5686–5692. doi: 10.1016/j.acra.2025.06.057.

76. Hadan, H., Wang, D. M., Mogavi, R. H., Tu, J., Zhang-Kennedy, L., & Nacke, L. E. (2024). The great AI witch hunt: Reviewers’ perception and (Mis)conception of generative AI in research writing. Computers in Human Behavior: Artificial Humans, 2(2), 100095. 10.1016/j.chbah.2024.100095

77. Liang, Weixin, et al. “GPT detectors are biased against non-native English writers.” Patterns 4.7 (2023).

78. Lingard L, Chandritilake M, de Heer M, Klasen J, Maulina F, Olmos-Vega F, St-Onge C. (Dec 2023) Will ChatGPT’s free language editing service level the playing field in science communication?: Insights from a collaborative project with non-native English scholars. Perspectives on Medical Education 12(1), 565–574.

79. Wondimagegn D, Whitehead CR, Cartmill C, Rodrigues E, Correia A, Lins TS, Costa MJ. Faster, higher, stronger–together? A bibliometric analysis of author distribution in top medical education journals. BMJ Global Health. 2023 Jun 15;8(6).

80. Maggio LA, Costello JA, Ninkov AB, Frank JR, Artino Jr AR. The voices of medical education scholarship: Describing the published landscape. Medical education. 2023 Mar;57(3):280–9.

81. Cheng A, Calhoun A, Reedy G. Artificial intelligence⍰assisted academic writing: recommendations for ethical use Advances in Simulation (2025) 10:22. 10.1186/s41077-025-00350-6

82. Ahmed S BaHammam (2025) The Transparency Paradox: Why Researchers Avoid Disclosing AI Assistance in Scientific Writing, Nature and Science of Sleep,, 2569–2574, DOI: 10.2147/NSS.S568375

83. Hartberg Y. AI Disclosures are A Joke. Chronicle of Higher Education. August 2025. Accessed online Nov 3, 2025. https://www.chronicle.com/article/ai-writing-disclosures-are-a-joke-heres-how-to-improve-them

84. Resnik DB & Hosseini M (24 Mar 2025). Disclosing artificial intelligence use in scientific research and publication: When should disclosure be mandatory, optional, or unnecessary? Accountability in Research, DOI: 10.1080/08989621.2025.2481949

85. Weaver, KD. The AI Disclosure (AID) Framework. C&RL News, 85(10), 407–411 (2024) https://arxiv.org/pdf/2408.01904

86. Maggio LA, Costello JA, Brown KR, Artino AR Jr, Durning SJ, Ma TL. Time to Publication in Medical Education Journals: An Analysis of Publication Timelines During COVID-19 (2019-2022). Perspect Med Educ. 2024 Oct 11;13(1):507–517. doi: 10.5334/pme.1287.

