## Supplemental Appendix A for "The Presence and Nature of AI-Use Disclosure Statements in Medical Education Journals: A bibliometric study"

### Appendix A – Frequency of AI-Use Disclosure Statements by Countries

| Country | Frequency |
|---|---|
| United States of America | 8 |
| Germany | 6 |
| Iran | 5 |
| Australia | 5 |
| Japan | 3 |
| China | 3 |
| Nepal | 2 |
| New Zealand | 2 |
| Saudi Arabia | 1 |
| Netherlands | 1 |
| Brazil | 1 |
| Canada | 1 |
| South Africa | 1 |
| Turkey | 1 |
| Bangladesh | 1 |
| Thailand | 1 |
| Hungary | 1 |
| Singapore | 1 |
| England | 1 |
| Pakistan | 1 |
| Palestine | 1 |
| Uruguay | 1 |
| Egypt | 1 |
| Qatar | 1 |
